# The COVID-19 burnout scale: Development and initial validation

**DOI:** 10.1101/2022.10.20.22281317

**Authors:** Petros Galanis, Aglaia Katsiroumpa, Panayota Sourtzi, Olga Siskou, Olympia Konstantakopoulou, Daphne Kaitelidou

## Abstract

We developed and validated a self-assessment instrument to measure COVID-19 pandemic-related burnout in the general population. We assessed the psychometric properties of the COVID-19 burnout scale (COVID-19-BS). Exploratory and confirmatory factor analysis identified three factors for the COVID-19-BS; emotional exhaustion, physical exhaustion, and exhaustion due to measures against the COVID-19. Cronbach’ s alpha coefficients for the three factors and the COVID-19-BS ranged from 0.860 to 0.921. Kaiser-Meyer-Olkin value was 0.945 and p-value for Bartlett test was <0.001 indicating highly acceptable values. Convergent validity results indicated a significant positive correlation between COVID-19-BS and anxiety and depression. Known-groups analysis identified the ability of COVID-19-BS to discriminate groups according to gender, chronic condition, and health status. Our findings indicate that the final 13-item model of COVID-19-BS is a brief, easy to administer, valid and reliable scale for assessing COVID-19-related burnout in the general public.

## Introduction

The COVID-19 pandemic has caused many psychological problems in general population including depression, fear, stress, anxiety, post-traumatic stress disorder, psychological distress, and insomnia (Luo et al., 2020; Wu et al., 2021; Xiong et al., 2020). A meta-analysis found that the overall prevalence of anxiety among the general population during the COVID-19 pandemic is 38.1%, the overall prevalence of depression is 34.3% and the overall prevalence of psychological distress is 37.5% (Necho et al., 2021). Main predictors of psychological problems are female gender, younger age, lower socioeconomic status, presence of a chronic condition, social isolation, and unemployment (Luo et al., 2020; Xiong et al., 2020).

Longitudinal studies revealed that stress, depression, and anxiety levels in general population remain high during the pandemic (Brailovskaia et al., 2021; Somma et al., 2021; Wang et al., 2020). Thus, we should consider the stress related to the pandemic as chronic and severe. Moreover, burnout could be defined as a psychological syndrome caused by excessive and prolonged stress that has not been successfully managed (Maslach & Leiter, 2016). Initially, burnout was identified as emotional exhaustion, depersonalization (feelings of cynicism), and lack of personal accomplishment in the workplace context (Maslach et al., 2001). However, burnout happens when an individual feels emotionally, mentally and physically exhausted, and incapable of meeting constant demands (Queen & Harding, 2020).

After a tumultuous three years of the pandemic, people feel exhausted and disappointed for a number of reasons; some people have lost their jobs, many have worked in the confines of home, parents have to balance childcare and personal need, some COVID-19 patients have to recover from long-term COVID effects, etc. (Queen & Harding, 2020). During the pandemic, several pandemic-related burnout/fatigue terms have been coined in the literature, e.g. pandemic burnout, societal (or social) pandemic burnout, pandemic fatigue, behavioral fatigue, quarantine fatigue etc. (Harvey, 2020; Petherick et al., 2021; Queen & Harding, 2020). Research on the pandemic-related burnout has focused on work-related burnout especially among healthcare workers ignoring burnout among the general population (Galanis et al., 2021; Ghahramani et al., 2021; Sharifi et al., 2021).

There are only two scales that measure pandemic-related burnout in the general population (Lau et al., 2022; Yıldırım & Solmaz, 2022). However, Yıldırım & Solmaz (2022) did not develop a new scale but they adapted and validated the Burnout Measure-Short Version in the case of the COVID-19 pandemic (Malach-Pines, 2005). Also, Lau et al. (2022) developed and validated a COVID-19 burnout frequency scale but this scale has been validated within the context of a dynamic zero-COVID-19 policy in Hong Kong. Therefore, these two scales have disadvantages and are not appropriate to measure the pandemic-related burnout of the general population in a global context.

Given that the COVID-19 pandemic is continuing to threaten mental health of individuals, and the scarcity of scales that directly measure pandemic-related burnout of the general population, we sought to develop a valid and reliable instrument to measure pandemic-related burnout in the general population.

## Methods

### Development of the scale

We took several steps to develop the scale items and achieve content validity (McCoach et al., 2013). First, we performed a literature review to identify questionnaires that measure work-related burnout among healthcare workers prior to the onset of pandemic and during the pandemic since scholars have mainly investigated occupational burnout among healthcare providers (Galanis et al., 2021; Ghahramani et al., 2021; Low et al., 2019; Sharifi et al., 2021; Woo et al., 2020). Also, we reviewed burnout literature regarding the COVID-19 pandemic, e.g. the policy framework for supporting pandemic prevention and management by the World Health Organization (World Health Organization, 2020).

Second, based on the literature review we developed items that could measure pandemic-related burnout in the general population. In that step, we developed 25 relevant items. All items were COVID-19 oriented (e.g., “I feel emotionally tired because of the COVID-19 pandemic”, “I feel emotionally tired because of the COVID-19 pandemic”, “I feel tired applying personal protection measures”, etc.).

Third, a panel of experts (n=10) from different fields and backgrounds (mental healthcare professionals, psychologists, sociologists, physicians, and nurses) reviewed the initial set of 25 items. In that step, we asked experts to evaluate how well each item corresponds to pandemic-related burnout in the general population. Experts rated each item as “essential”, “useful but not essential” or “not essential”. Then we calculated the content validity ratio for each item using the below formula:

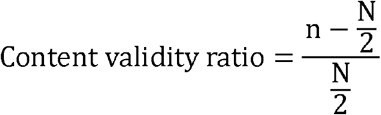

In this formula, “N” is the total number of experts (10 in our study) and “n” is the number of experts who rate an item as “essential”. Items with a value greater than 0.80 were considered as appropriate and therefore remained (Ayre & Scally, 2014). We removed the following eight items due to unsatisfactory content validity ratio: “I feel anxiety because I have to comply with personal preventive measures”, “I am afraid there may be another lockdown”, “I feel fear because of the COVID-19 pandemic”, “I feel trapped because of the COVID-19 pandemic”, “I am afraid to travel because of the COVID-19 pandemic”, “I am confused about how to comply with personal preventive measures, e.g. when and where to wear the facing mask, when to do a rapid test, a PCR test, etc.”, “I feel tired of taking preventive measures against the COVID-19, e.g. taking vitamins, etc.”, and “I feel tired of the reduced effectiveness of the measures against the pandemic”.

Fourth, we conducted cognitive interviews with 10 participants from the general population to assess face validity and gain insights into participants understanding of 17 scale items (Meadows, 2021). All respondents understood the item descriptions, and we only made minor revisions in three items in order to improve the understanding of the scale. In particular, respondents asked for examples in three items.

After all, the scale included 17 items. All items are negatively formulated in order to improve the understanding of the scale (e.g., “I feel emotionally tired because of the COVID-19 pandemic”). Each item is rated on a five-point Likert scale (1-5): strongly disagree (1), disagree (2), undecided (3), agree (4), and strongly agree (5). Higher values in all items indicate higher levels of burnout in the general population.

### Procedure

We created the COVID-19 burnout scale (COVID-19-BS) in Greek. All adults aged 18 years or older that were being able to read and understand Greek could participate in our study. We used Google forms to create an anonymous online version of the scale and we disseminated it through social media and e-mail. Thus, a convenience sample was obtained. Data were collected during September 2022.

### Measures

#### Demographic data

We collected demographic data of the participants including gender, age, chronic condition, self-assessment of health status, and history of COVID-19 infection.

#### Patient Health Questionnaire-4 (PHQ-4)

The PHQ-4 consists of four items assessing anxiety and depression (Kroenke et al., 2009). A sample item is the following: “Over the last two weeks, how often have you been bothered by the following problem? Feeling nervous, anxious or on edge”. Each item is rated on a four-point Likert scale from 0 (not at all) to 3 (nearly every day). The overall PHQ-4 score is estimated by adding all four items and such that overall score can range from 0 to 12. Higher overall PHQ-4 score indicates greater anxiety and depression. PHQ-4 has been validated in Greek language (Karekla et al., 2012). Cronbach’ s alpha for PHQ-4 was 0.870 in our study.

#### Single Item Burnout measure (SIB)

The SIB measure asking respondents to rate their level of burnout on a scale from zero (not at all burnt out) to 10 (extremely burnt out) and seems to be an easy and valid screening measure of burnout (Hansen & Pit, 2016).

### Participants

Study sample included 1256 participants (69.9% females; age ranged from 18 to 80 with a mean age of 39.15 years, standard deviation = 11.87) from the general public in Greece. Of the participants, 19.4% reported a history of chronic condition, while 69.4% reported confirmed history of COVID-19 infection. Demographic characteristics of participants are shown in Table 1.

**Table 1.**
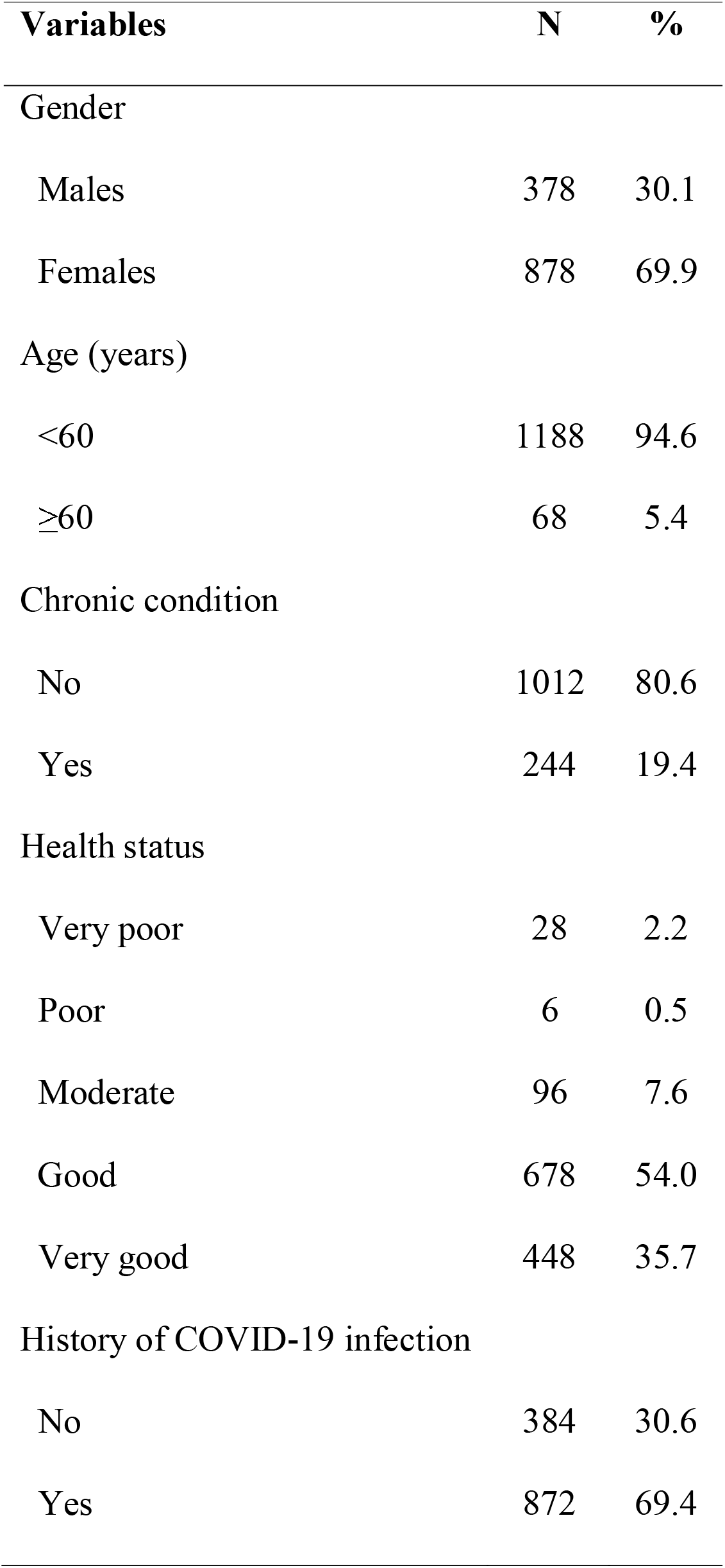
Demographic characteristics of participants (n=1256).

| **Variables** | **N** | **%** |
| --- | --- | --- |
| **Gender** |  |  |
| Males | 378 | 30.1 |
| Females | 878 | 69.9 |
| **Age (years)** |  |  |
| <60 | 1188 | 94.6 |
| ≥60 | 68 | 5.4 |
| **Chronic condition** |  |  |
| No | 1012 | 80.6 |
| Yes | 244 | 19.4 |
| **Health status** |  |  |
| Very poor | 28 | 2.2 |
| Poor | 6 | 0.5 |
| Moderate | 96 | 7.6 |
| Good | 678 | 54.0 |
| Very good | 448 | 35.7 |
| **History of COVID-19 infection** |  |  |
| No | 384 | 30.6 |
| Yes | 872 | 69.4 |

### Ethics

We did not ask about personal data and participation was anonymous and voluntary. We informed participants about the objective of the study and we obtained informed consent electronically before data collection. We followed the guidelines of the Declaration of Helsinki to conduct our study (World Medical Association, 2013). Study protocol was approved by the Ethics Committee of the Faculty of Nursing, National and Kapodistrian University of Athens (approval number; 370, 02-09-2021).

### Statistical analysis

#### Validity analysis

We conducted exploratory and confirmatory factor analysis to investigate construct validity of the COVID-19-BS. We randomly divided the participants into two subsamples (n=628 for the first subsample and n=628 for the second subsample) to test the structure of the scale. First, we conducted exploratory factor analysis (EFA) on the first subsample to identify the underlying factor structure of the scale and then we performed confirmatory factor analysis (CFA) to verify the factor structure that emerged from the EFA. The cut-off criteria for the applicability of the EFA were the Kaiser-Meyer-Olkin (KMO) index >0.80, and the p-value for the Bartlett sphericity test <0.05. We applied varimax rotation method to extract factors in EFA using the following criteria: eigenvalues >1, factor loadings >0.60, communalities >0.40, and the total variance explained by the factors. We removed items with low communalities and/or low cross loadings, e.g. loading in more than one of the factors. Then, we performed CFA on the second subsample to examine the adequacy of fit of the factor model derived from the EFA. We presented several goodness of fit indices in CFA to assess overall model fit: chi-square divided by degree of freedom (x^2^/df), root mean square error of approximation (RMSEA), goodness of fit index (GFI), adjusted goodness of fit index (AGFI), Tucker-Lewis index (TLI), incremental fit index (IFI), normed fit index (NFI), and comparative fit index (CFI). Based on the recommended cut-off values, we considered x^2^/df < 5, RMSEA < 0.10, GFI, AGFI, TLI, IFI, NFI, CFI > 0.90 as acceptable fit indices. Also, we considered x^2^/df < 3, RMSEA < 0.08, and GFI, AGFI, TLI, IFI, NFI, CFI > 0.95 as good fit indices (Bentler et al., 1980; Hooper et al., 2008; Hu & Bentler, 1998; Steiger, 2007). Moreover, we examined the modification index and factor loadings. We removed items with large error correlations and cross-factor loadings. We conducted CFA using AMOS (version 23).

For each factor that emerged from the factor analysis we calculated the total score by dividing the sum of all answers in one factor with the total number of items included in this factor. In the same way, we calculated a total score for the COVID-19-BS.

Thus, the total score for each factor and the COVID-19-BS ranged from 1 to 5 with higher values indicating higher levels of burnout.

Subsequent validity analyses were performed on overall sample. We used the PHQ-4 and the SIB measure to assess the convergent validity of the COVID-19-BS. In particular, we expected a positive relationship between COVID-19-BS and PHQ-4 and SIB measure. We used Pearson’ s correlation coefficient to estimate the relationship between the COVID-19-BS factors and the PHQ-4 and the SIB measure since the scores in all scales followed the normal distribution.

To further evaluate the validity of the COVID-19-BS we applied the known-groups method in order to check the ability of the scale to discriminate females from males, older participants from younger participants, participants with a chronic condition from healthy participants, participants with a better health status from participants with a worse health status, and participants with a history of COVID-19 infection from participants without a history of COVID-19 infection. We measured health status in a five-point Likert scale: very poor, poor, moderate, good, and very good. We treated age and health status as dichotomous variables. In particular, we used the cut-off point of 60 years for age. Regarding health status, we merged the answers “very poor”, “poor”, “moderate”, and “good” in one category. We used independent samples t-test to estimate the differences between the total burnout score and demographic variables since the scores followed the normal distribution.

#### Reliability analysis

We used test-retest reliability, split-half reliability, and Cronbach’ s alpha to estimate reliability of the COVID-19-BS. Fifty participants completed the COVID-19-BS twice in five days in order to estimate test-retest reliability. In that case, we calculated Spearman’ s correlation coefficients for the 17 items of the COVID-19-BS in the two measurements since items were measured in an ordinal scale.

Also, we used the overall sample to perform split-half reliability and estimate Cronbach’ s alpha. In particular, we split the COVID-19-BS into two halves; one half composed of even-numbered items (n=6), while the other half composed of odd-numbered items (n=7). We calculated the total score for the two halves and then we calculated the Pearson’ s correlation coefficient between the two halves since the scores followed the normal distribution. Moreover, we measured Cronbach’ s alpha for the COVID-19-BS and the factors in order to estimate the internal consistency of the scale. Cronbach’ s alpha values greater than 0.7 were considered as acceptable (Hair et al., 2017).

Finally, we calculated corrected item-total correlation for items included in a factor. Values greater than 0.30 are acceptable for item-total correlation (Nunnally & Bernstein, 1994).

All statistical tests were two sided and p-values less than 0.05 were considered as statistically significant. We used IBM SPSS 21.0 (IBM Corp. Released 2012. IBM SPSS Statistics for Windows, Version 21.0. Armonk, NY: IBM Corp.) for statistical analysis.

## Results

### Validity analysis

#### Factor analysis

Using the first subsample, an EFA was conducted. Kaiser-Meyer-Olkin value was 0.945 and p-value for Bartlett test was <0.001 indicating that the sample size was adequate to perform EFA. Based on communalities and cross loading, we removed the following items: “I have lost my hope that the COVID-19 pandemic will end soon”, “I feel anxiety because of the COVID-19 pandemic”, “I feel nervous because of the COVID-19 pandemic”, and “I feel helpless to deal with the COVID-19 pandemic”. Thus a 13-items model was emerged that explained 71.573% of the variance of the COVID-19-BS (Table 2). EFA identified three factors for the COVID-19-BS; emotional exhaustion (five items), physical exhaustion (four items), and exhaustion due to measures against the COVID-19 (four items).

**Table 2.**
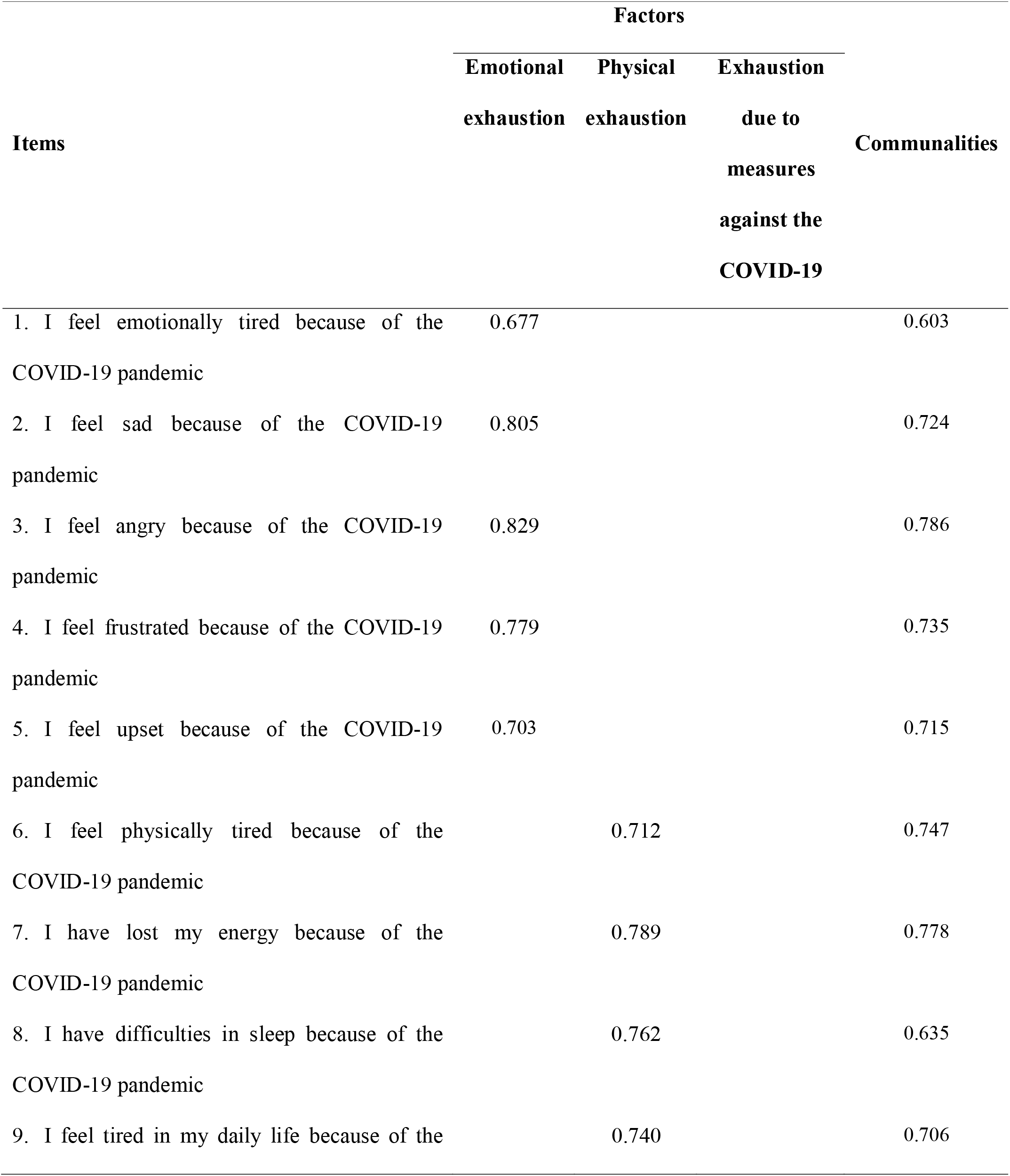
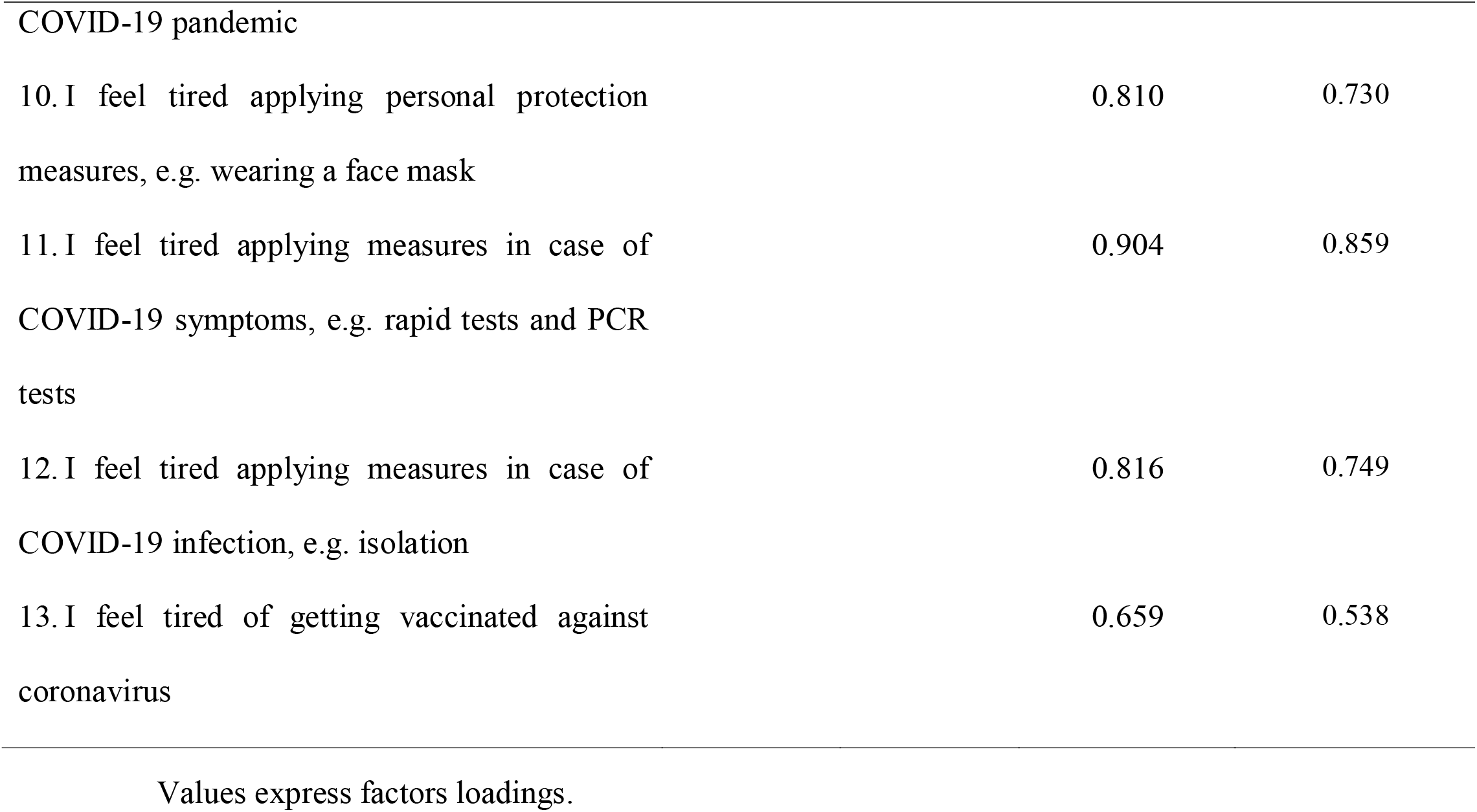
Exploratory factor analysis for the COVID-19 burnout scale (n=628).

Then, we conducted a CFA using the second subsample. The construction of the CFA model was based on the factor structure generated by EFA. The goodness-of-fit statistics suggested that the 13-item model of the COVID-19-BS without errors correlation provided an acceptable fit to data according to RMSEA, GFI, TLI, IFI, NFI, and CFI but a poor fit to data according to x^2^/df and AGFI (Table 3). Taken into account the error correlations, the model significantly improved. In particular, now the 13-item model of the COVID-19-BS provided a good fit to data according to RMSEA, IFI, and CFI, and an acceptable fit to data according to x^2^/df, GFI, AGFI, TLI, and NFI (Table 3). Moreover, the standardized regression weights between 13 items and three factors were statistically significant and ranged from 0.721 to 0.859. Also, the correlations between the three factors were positive (from 0.532 to 0.812) and statistically significant. Since the correlation between the factors “emotional exhaustion” and “physical exhaustion” was high we merged these two factors and we performed a CFA. The results, as shown in Table 3, were considerably worse than the three factor model. The path diagram of 13-item CFA model is shown in Figure 2.

**Table 3.** Goodness-of-fit statistics of the different confirmatory factor analysis

| **Models** | **$\chi^2$** | **Df** | **$\chi^2/df$** | **RMSEA** | **GFI** | **AGFI** | **TLI** | **IFI** | **NFI** | **CFI** |
| --- | --- | --- | --- | --- | --- | --- | --- | --- | --- | --- |
| 13-item model with<br>three factors without<br>errors correlation | 425.966 | 67 | 6.300 | 0.092 | 0.911 | 0.880 | 0.922 | 0.933 | 0.922 | 0.933 |
| 13-item model with<br>three factors included<br>errors correlation | 293.814 | 60 | 4.897 | 0.079 | 0.935 | 0.902 | 0.943 | 0.957 | 0.946 | 0.956 |
| 13-item model with<br>two factors | 919.600 | 75 | 12.261 | 0.134 | 0.801 | 0.759 | 0.836 | 0.843 | 0.831 | 0.843 |

**Figure 1.**
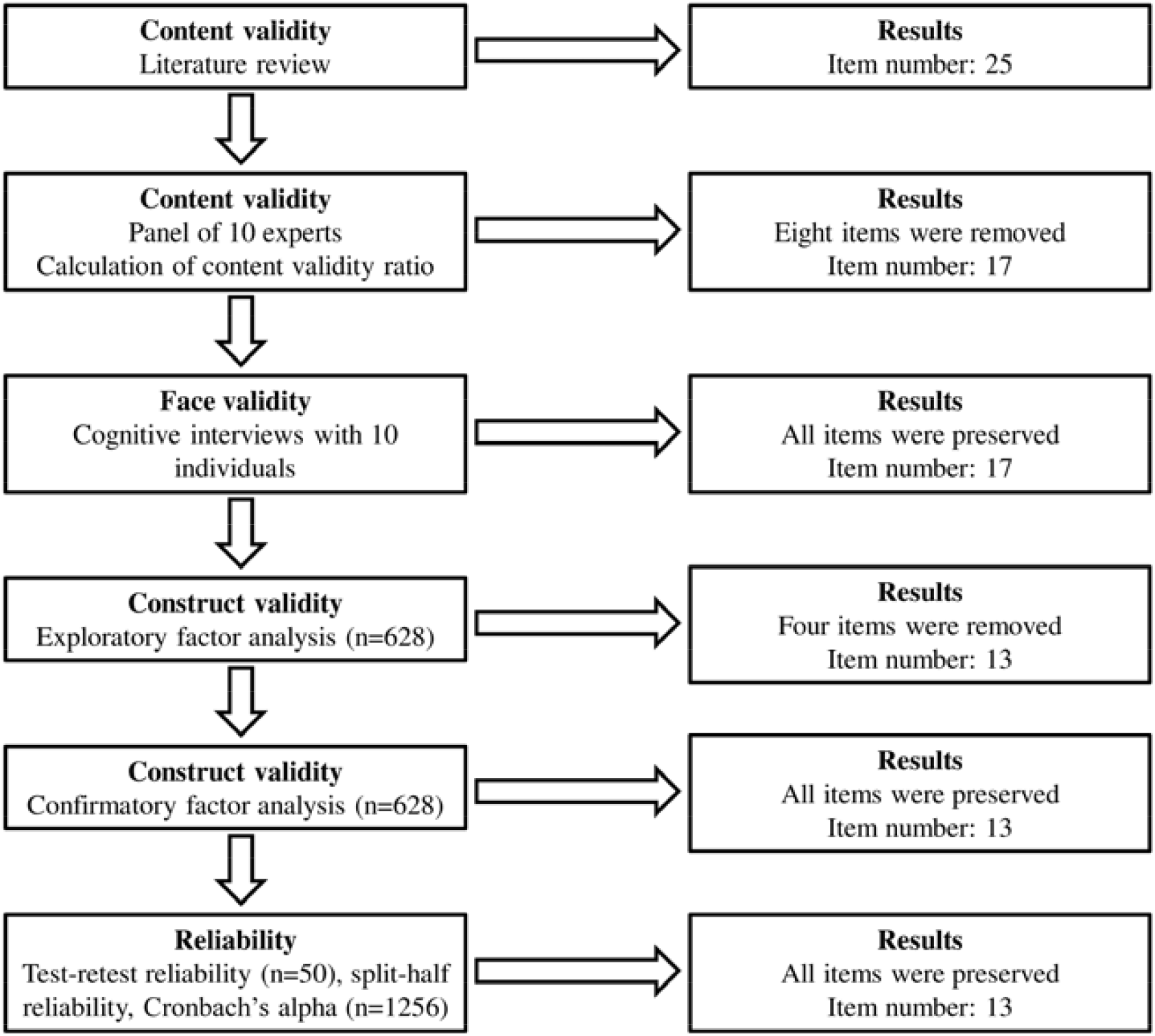
Development and validation of the COVID-19 burnout scale.

**Figure 2.**
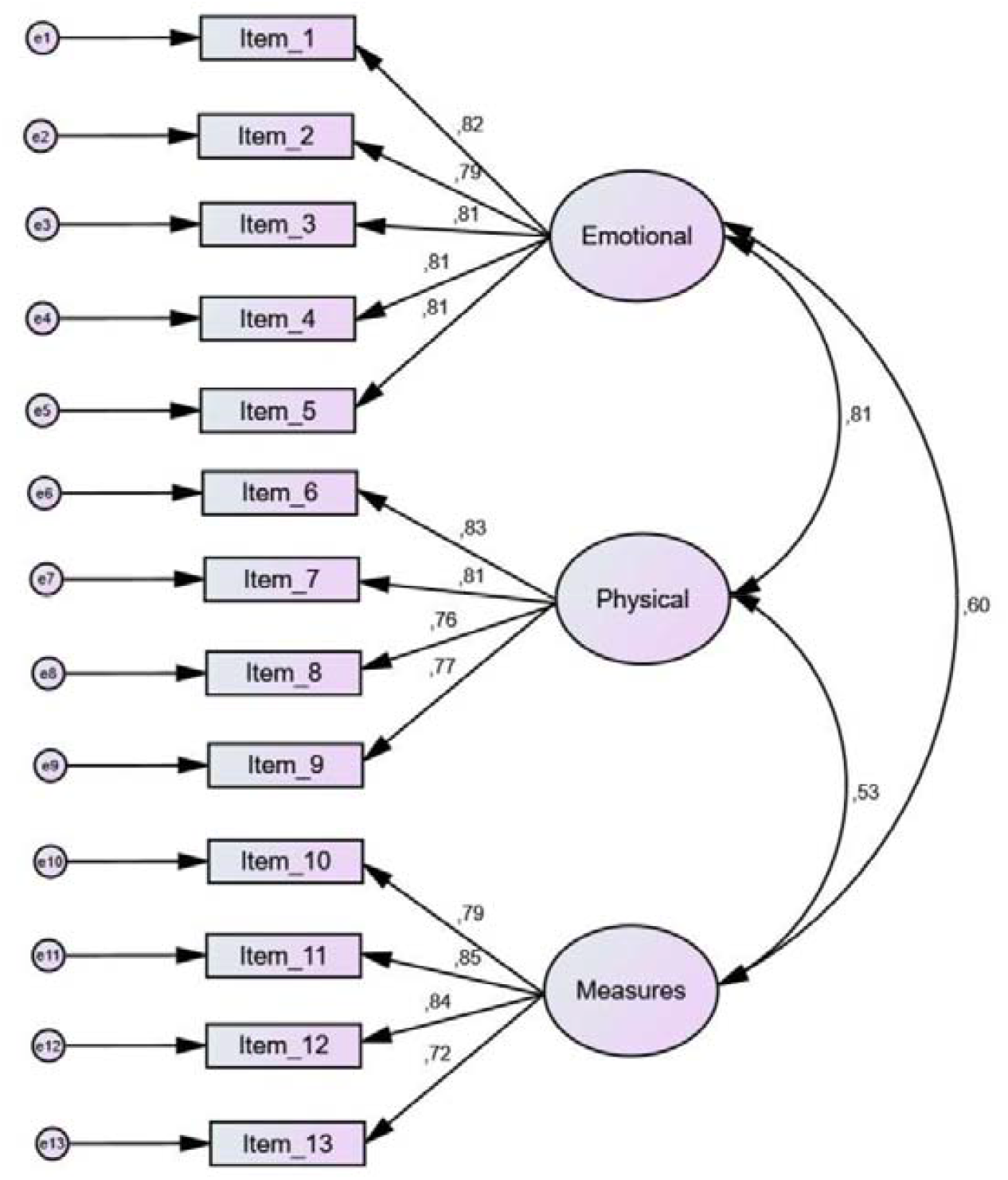
Confirmatory factor analysis of COVID-19 burnout scale and regression/correlation values.

Finally, based on the results of exploratory and confirmatory factor analysis, we concluded that a three-factor model including 13 items was appropriate for the COVID-19-BS; emotional exhaustion (five items), physical exhaustion (four items), and exhaustion due to measures against the COVID-19 (four items), (Supplementary Table 1).

#### Convergent validity

Pearson’ s correlation coefficients between the PHQ-4 and the factors of the COVID-19-BS ranged from 0.379 to 0.514 (p<0.001 in all cases) suggesting that individuals with higher levels of depression and anxiety may also had higher levels of burnout, or vice versa (Table 4). Moreover, Pearson’ s correlation coefficients between the SIB measure and the factors of the COVID-19-BS ranged from 0.253 to 0.430 (p<0.001 in all cases). Therefore, the convergent validity of the COVID-19-BS was very good.

**Table 4.** Pearson’ s correlation coefficient between the COVID-19 burnout scale and the PHQ-4 and the SIB measure (n=1256).

| COVID-19 burnout scale | PHQ-4 |  | SIB measure |  |
| --- | --- | --- | --- | --- |
|  | Pearson's correlation | P-value | Pearson's correlation | P-value |
|  | coefficient |  | coefficient |  |
| Emotional exhaustion | 0.445 | <0.001 | 0.330 | <0.001 |
| Physical exhaustion | 0.514 | <0.001 | 0.430 | <0.001 |
| Exhaustion due to measures<br>against the COVID-19 | 0.379 | <0.001 | 0.253 | <0.001 |
| Total scale | 0.524 | <0.001 | 0.395 | <0.001 |

**Table 5.** Total burnout score according to demographic characteristics of participants (n=1256).

| **Variables** | **Mean total burnout score** | **Standard deviation** | **P-value<sup>a</sup>** |
| --- | --- | --- | --- |
| Gender |  |  | <0.001 |
| Males | 2.671 | 0.920 |  |
| Females | 3.108 | 0.983 |  |
| Age (years) |  |  | 0.068 |
| <60 | 2.990 | 0.976 |  |
| ≥60 | 2.738 | 1.102 |  |
| Chronic condition |  |  | <0.001 |
| No | 2.933 | 0.966 |  |
| Yes | 3.158 | 1.039 |  |
| Health status |  |  | <0.001 |
| Very poor to good | 3.116 | 0.999 |  |
| Very good | 2.725 | 0.904 |  |
| History of COVID-19 infection |  |  | 0.107 |
| No | 2.909 | 1.017 |  |
| Yes | 3.006 | 0.969 |  |

<sup>a</sup> Independent samples t-test

#### Known-groups validity

Levels of burnout was higher among females, participants with a chronic condition, and those with worse health status (p<0.001 in all cases) indicating the ability of the COVID-19-BS to discriminate different groups. We did not find relationship between burnout and age (p=0.068) and history of COVID-19 infection (p=0.107).

### Reliability analysis

We found an excellent reliability of the COVID-19-BS using the test-retest method. In particular, Spearman’ s correlation coefficient for the 17 items between the two measurements was >0.7 (p<0.05 for all items). Cronbach’ s alpha coefficient for the final 13-item model of COVID-19-BS was 0.921, while for the factor “emotional exhaustion” was 0.904, for the factor “physical exhaustion” was 0.860, and for the factor “exhaustion due to measures against the COVID-19” was 0.862. Factor loadings ranged from 0.659 to 0.904, while communalities ranged from 0.603 to 0.859. Regarding split-half reliability, Pearson’ s correlation coefficient between the two halves of the COVID-19-BS was very high (0.92, p<0.001). Corrected item-total correlation for items ranged from 0.543 to 0.749 (Supplementary Table 2). All correlation coefficients were statistically significant at p<0.05 level. Therefore, the reliability of the COVID-19-BS was excellent.

## Discussion

COVID-19-related burnout is a key concern among the general public but research until now has investigated only burnout among healthcare workers. Surprisingly, to date, COVID-19-related burnout in the general population has remained unknown. Only Lau et al. (2022) developed and validated a COVID-19 burnout frequency scale but their study was performed within the context of a dynamic zero-COVID-19 policy with strict measures. Thus, we developed and validated a scale to measure burnout of the general population within the context of pandemic. In our study, we created the COVID-19-BS and we evaluated the validity and the reliability of the scale with a large sample of the general population in Greece.

We found that COVID-19-BS has very good validity and reliability in evaluating burnout caused by the COVID-19 pandemic among general public. Thus, the COVID-19-BS is a specific tool for the assessment of burnout that it can accurately evaluate the burnout level of the individuals. Moreover, the COVID-19-BS is relatively brief since it consists of 13 items. Therefore, the COVID-19-BS is highly practical and could be used as a routine screening tool for assessment of individuals in everyday clinical care practice. In addition, the COVID-19-BS contains emotional exhaustion, physical exhaustion, and exhaustion due to measures against the COVID-19 subscales covering different aspects of burnout.

The results of our study showed that the final 13-item CFA model-to-data fit was adequate with standardized regression weights ranging from 0.721 to 0.889 and factor loadings ranging from 0.659 to 0.904. The COVID-19-BS also had a very high internal consistency as indicated by the Cronbach’ s alpha coefficient (all values were above 0.860). Moreover, convergent and known-groups validity analysis confirmed that the COVID-19-BS is a valid tool for measuring burnout in the general population.

COVID-19-BS demonstrated multi-dimensional structure since it consisted of three factors that can measure the level of burnout associated with emotional and physical exhaustion, and exhaustion due to measures against the COVID-19. Our three-factor model confirm the fact that exhaustion is the core aspect of burnout (Maslach & Leiter, 2016). Moreover, evidence shows that the adherence of individuals to COVID-19 preventive measures (e.g. face covering and social distancing) disrupts verbal and non-verbal communication and affects negatively wellbeing and quality of life (Oosthuizen et al., 2022).

We found a significant correlation between the COVID-19-BS and levels of depression and anxiety. Although there is a lack of studies searching the relationship between burnout and depression and anxiety in the general population during the pandemic, several studies support the correlation between these variables among healthcare workers, teachers, police officers and students (Chowdhury et al., 2022; Ghio et al., 2021; Hung & Liu, 2022; Karakose et al., 2022; Tehrani, 2022; Tomaszek & Muchacka-Cymerman, 2022; Ulfa et al., 2022). In addition, Yıldırım and Solmaz (2022) found that COVID-19 stress predicted COVID-19 burnout in a sample of Turkish adults (Yıldırım & Solmaz, 2022). Also, a recent meta-analysis including studies before the pandemic found a significant association between burnout and depression and anxiety (Koutsimani et al., 2019).

Known-groups method identified that females, participants with a chronic condition, and those with worse health status experienced higher levels of burnout. Several systematic reviews confirm that female gender is associated with high prevalence of burnout among healthcare workers during the pandemic (Claponea et al., 2022; Lluch et al., 2022; Shaikh et al., 2022). Moreover, several studies during the pandemic confirmed that poor physical/mental health was a predictor for increased healthcare workers’ burnout (Arrogante & Aparicio-Zaldivar, 2020; Kim et al., 2022; La Torre et al., 2021; Malagón-Aguilera et al., 2020).

### Limitations

Our study has some limitations that should be taken into consideration. Selection bias is probable in our study since we used a convenience sample of the general population. For instance, considering the large proportion of the participants being female and young, our sample may not represent the general population. Moreover, we developed and validated our scale in a Greek cultural context. Thus, further studies with random samples should be conducted in order to evaluate the validity and reliability of the COVID-19-BS. Moreover, future research should evaluate the applicability of the COVID-19-BS to different populations and cultural settings. In addition, longitudinal studies should examine how burnout changes over time in response to the development of the pandemic. We examined the convergent validity of the COVID-19-BS using two constructs (PHQ-4 and SIB measure). There is a need to examine the association between the COVID-19-BS and other constructs. Finally, we used five demographic characteristics to evaluate the discrimination validity of the COVID-19-BS. Future studies could use more socio-demographic variables to establish the validity of the scale.

## Conclusions

Findings from this study support using the COVID-19-BS as a brief, easy to administer, valid and reliable scale for assessing COVID-19-related burnout in the general public. Despite the limitations mentioned above, the COVID-19-BS could be used as a timely screening tool to assess individuals’ burnout and to identify vulnerable groups. Thanks to its very good psychometric properties, the COVID-19-BS may help clinicians, health educators, scholars and policy makers to recognize the high-risk groups for COVID-19 prevention and education.

**Supplementary Table 1.** Final structure of the 13-item model of the COVID-19 burnout scale.

| **Factors** | **Items** |
| --- | --- |
| **Emotional exhaustion** | 1. I feel emotionally tired because of the COVID-19 pandemic |
|  | 2. I feel sad because of the COVID-19 pandemic |
|  | 3. I feel angry because of the COVID-19 pandemic |
|  | 4. I feel frustrated because of the COVID-19 pandemic |
|  | 5. I feel upset because of the COVID-19 pandemic |
| **Physical exhaustion** | 6. I feel physically tired because of the COVID-19 pandemic |
|  | 7. I have lost my energy because of the COVID-19 pandemic |
|  | 8. I have difficulties in sleep because of the COVID-19 pandemic |
|  | 9. I feel tired in my daily life because of the COVID-19 pandemic |
| **Exhaustion due to measures against the COVID-19** | 10. I feel tired applying personal protection measures, e.g. wearing a face mask |
|  | 11. I feel tired applying measures in case of COVID-19 symptoms, e.g. rapid tests and PCR tests |
|  | 12. I feel tired applying measures in case of COVID-19 infection, e.g. isolation |
|  | 13. I feel tired of getting vaccinated against coronavirus |

**Supplementary Table 2.** Corrected item-total correlation for the 13-item model of the COVID-19 burnout scale.

| **Factors** | **Items** | **Corrected item-total correlations** |
| --- | --- | --- |
| **Emotional exhaustion** | 1. I feel emotionally tired because of the COVID-19 pandemic | 0.656 |
|  | 2. I feel sad because of the COVID-19 pandemic | 0.634 |
|  | 3. I feel angry because of the COVID-19 pandemic | 0.695 |
|  | 4. I feel frustrated because of the COVID-19 pandemic | 0.715 |
|  | 5. I feel upset because of the COVID-19 pandemic | 0.749 |
| **Physical exhaustion** | 6. I feel physically tired because of the COVID-19 pandemic | 0.720 |
|  | 7. I have lost my energy because of the COVID-19 pandemic | 0.697 |
|  | 8. I have difficulties in sleep because of the COVID-19 pandemic | 0.543 |
|  | 9. I feel tired in my daily life because of the COVID-19 pandemic | 0.678 |
| **Exhaustion due to measures against the COVID-19** | 10. I feel tired applying personal protection measures, e.g. wearing a face mask | 0.609 |
|  | 11. I feel tired applying measures in case of COVID-19 symptoms, e.g. rapid tests and PCR tests | 0.598 |
|  | 12. I feel tired applying measures in case of COVID-19 infection, e.g. isolation | 0.629 |
|  | 13. I feel tired of getting vaccinated against coronavirus | 0.564 |

## Data Availability

All data produced in the present study are available upon reasonable request to the authors

## Declarations

### Funding

None

### Competing interests

None

### Ethical approval

All procedures followed were in accordance with the ethical standards of the responsible committee on human experimentation (institutional and national) and with the Helsinki Declaration of 1975, as revised in 2000. Informed consent was obtained from all participants for being included in the study.

### Conflict of interest

All authors declare that they have no conflict of interest.

## Notes

### Competing Interest Statement

The authors have declared no competing interest.

### Funding Statement

This study did not receive any funding

### Author Declarations

Study protocol was approved by the Ethics Committee of the Faculty of Nursing, National and Kapodistrian University of Athens (approval number; 370, 02-09-2021).

